# Heterogeneity in risk, testing and outcome of COVID-19 across outbreak settings in the Greater Toronto Area, Canada: an observational study

**DOI:** 10.1101/2020.06.12.20129783

**Authors:** Linwei Wang, Huiting Ma, Kristy C.Y. Yiu, Andrew Calzavara, David Landsman, Linh Luong, Adrienne K. Chan, Rafal Kustra, Jeffrey C Kwong, Marie-Claude Boily, Stephen Hwang, Sharon Straus, Stefan D Baral, Sharmistha Mishra

**Affiliations:** MAP Centre for Urban Health Solutions, St. Michael’s Hospital, University of Toronto, Toronto, Canada; ICES, Toronto, Canada; Division of Infectious Diseases, Department of Medicine, University of Toronto, Toronto, Canada; Division of Infectious Diseases, Sunnybrook Health Sciences, University of Toronto, Toronto, Canada; Dalla Lana School of Public Health, University of Toronto, Toronto, Canada; Department of Family and Community Medicine, Faculty of Medicine, University of Toronto, Toronto, Canada; Department of Infectious Disease Epidemiology, Faculty of Medicine, Imperial College, London, United Kingdom; Division of General Internal Medicine, Department of Medicine, University of Toronto, Toronto, Canada; Department of Medicine, St. Michael’s Hospital, University of Toronto, Toronto, Canada; Bloomberg School of Public Health, Johns Hopkins University, Baltimore, MD, United States

## Abstract

**Background:** We compared the risk of, testing for, and death following COVID-19 infection across three settings (long-term care homes (LTCH), shelters, the rest of the population) in the Greater Toronto Area (GTA), Canada.

**Methods:** We sourced person-level data from COVID-19 surveillance and reporting systems in Ontario, and examined settings with population-specific denominators (LTCH residents, shelters, and the rest of the population). We calculated cumulatively, the diagnosed cases per capita, proportion tested for COVID-19, daily and cumulative positivity, and case fatality proportion. We estimated the age- and sex-adjusted relative rate ratios for test positivity and case fatality using quasi-Poisson regression.

**Results:** Between 01/23/2020-05/25/2020, we observed a shift in the proportion of cases: from travel-related and into LTCH and shelters. Cumulatively, compared to the rest of the population, the number of diagnosed cases per 100,000 was 59-fold and 18-fold higher among LTCH and shelter residents, respectively. By 05/25/2020, 77.2% of LTCH residents compared to 2.4% of the rest of the population had been tested. After adjusting for age and sex, LTCH residents were 2.5 times (95% confidence interval (CI): 2.3-2.8) more likely to test positive. Case fatality was 26.3% (915/3485), 0.7% (3/402), and 3.6% (506/14133) among LTCH residents, shelter population, and others in the GTA, respectively. After adjusting for age and sex, case fatality was 1.4-fold (95%CI: 1.1-1.9) higher among LTCH residents than the rest of the population.

**Interpretation:** Heterogeneity across micro-epidemics among specific populations in specific settings may reflect underlying heterogeneity in transmission risks, necessitating setting-specific COVID-19 prevention and mitigation strategies.

## Introduction

Globally as of May 25, 2020, 5.4 million people have been diagnosed with COVID-19, of whom 345,000 have died (1). In Canada, 85,0000 people have been diagnosed with COVID-19, including 6,000 people who have died (1). Across North America, large urban centres such as the Greater Toronto Area (GTA) have borne the highest burden of cases (2). By May 25, 18,020 cases of COVID-19 had been diagnosed in the GTA’s population of six million people (2), accounting for over two-thirds of all cases in Ontario, and 21.4% of national cases (1, 2).

Congregate living settings have been disproportionately affected in past respiratory outbreaks (3). Settings such as long term care homes (LTCH) and homeless shelters are vulnerable partly due to design barriers (e.g. shared living quarters and communal spaces) to physical distancing, and under-resourcing of infection prevention and control measures (4-8). COVID-19 outbreaks in congregate settings have been similarly observed globally (9) and across Canada (10-13). For example, 416 of 630 LTCH in Ontario, excluding retirement homes, have had a COVID-19 outbreak (12), and 14 of 53 shelters in the City of Toronto are in active outbreak as of June 4^th^ (13).

Lessons learned from past epidemics suggest that disproportionate risks across settings contribute to the spread and outcomes of infection (14). Thus, a key feature of an epidemic response is quantifying heterogeneity in ‘what has happened’ with respect to disproportionate risks, a process often referred to as an epidemic appraisal (15, 16). We characterized, using the best available data sources, heterogeneity over time in testing (proportion tested), risk (diagnosed cases per capita, testing positivity), and outcome (death) following COVID-19 diagnosis in the GTA across three settings with available population-size data: LTCH residents, persons using shelters, and the rest of the population.

## Methods

### Study setting

We defined the GTA as the City of Toronto, York, Peel, Halton and Durham public health units (13, 17-20).

### Data sources

We sourced COVID-19 surveillance and reporting systems for information on the number of laboratory-confirmed cases (2), testing and results (21), and death (2). Data were obtained through May 25, 2020, four months after the first case of COVID-19 in the GTA (January 23)(2). We used person-level data in our analyses (2, 21); and cross-checked estimates for LTCH residents against aggregate data from the provincial LTCH tracker which reports a daily census of LTCH residents with confirmed and probable COVID-19 and deaths (12).

### The integrated Public Health Information System (iPHIS)

iPHIS is the provincial reportable diseases of confirmed and probable cases (2). Each public health unit submits person-level data to iPHIS, including the date of case report to public health; outcomes (e.g. death); case acquisition; demographics (sex, age) (2); and an outbreak-ID if the case was related to an outbreak in congregate settings (22, 23). A subset of iPHIS data (confirmed cases only; age-group instead of age in years) were made available by the Ontario Ministry of Health to the Ontario Modeling Consensus Table and used for these analyses.

### Ontario Laboratories Information System (OLIS)

OLIS contains data on COVID-19 tests submitted from hospitals, commercial laboratories, the provincial public health laboratory, and including testing at the COVID-19 assessment centres (21). OLIS includes test-level (date, result), and person-level (sex, age, address) data. Patient addresses were used to classify cases in the GTA and residents of LTCH. OLIS data were linked to records of emergency department visits and hospital admissions. Individuals with a record of emergency department visit or hospital admission in the past year and with an ‘homelessness’ indicator at the time of the service were identified as persons who may use shelters (24). These datasets were linked using unique encoded identifiers and analyzed at ICES (25).

Cross-checks identified the following: as of May 25, OLIS captured 90%, 96%, and 24% of confirmed cases in the GTA among overall population, LTCH residents, and persons using shelters, respectively, compared to iPHIS data (**Appendix-1 Figure-1A-C**). iPHIS recorded 915 deaths among LTCH residents with laboratory-confirmed COVID-19 versus 1032 recorded in the provincial LTCH tracker (**Appendix-1 Figure-1D**); the latter includes residents diagnosed with COVID-19 at time of death and thus may not yet be recorded in iPHIS.

### Population-specific denominators

We estimated the population of LTCH residents using the total LTCH bed capacity in the GTA, assuming complete occupancy (12). The population denominator for persons using shelters was sourced from the literature (26-33) (**Appendix-2**). To estimate the size of the rest of the population, we subtracted the above estimates from census-derived GTA population size (34).

### Statistical analyses

First, we calculated the cumulative and daily number and proportion of diagnosed cases over time in mutually exclusive categories in iPHIS: congregate settings (LTCH residents, staff, or other [e.g., volunteers], shelters, other congregate outbreak settings [hospitals, correctional facilities, retirement homes, group homes, and others not yet classified such as workplaces]; travel-related; and community settings (with versus without epidemiological link). Cases with missing information on setting excluded congregate settings.

Second, we examined differences in the three settings for which we had data on the population size (LTCH residents, persons using shelters, and the rest of the population): cumulative diagnosed cases per capita, cumulative proportion of population tested for COVID-19, daily and cumulative proportion of tests that were positive, and the cumulative case fatality proportion.

Third, we compared the age and sex distributions of diagnosed cases, proportion tested for COVID-19, and case fatality across the three settings using chi-squared tests. We used quasi-Poisson regression models (35) to estimate test positivity rate ratio, and case fatality rate ratio with 95% confidence intervals (CIs) among LTCH residents, and persons using shelters, separately, compared to the rest of the population; and adjusting for age (<50/50-59/60-69/70-70/80 years and older) and sex.

We used R version 3.6.3 (36) for data cleaning and analyses.

## Results

### Distribution in diagnoses over time across settings

By May 25, there were 18,020 diagnosed cases (263 cases per 100,000 population) in the GTA (**Figure-1A)**, with 250-350 newly diagnosed cases per day in the week prior to May 25 (**Appendix-1 Figure-2A)**. Diagnosed cases with a known travel history accounted for more than 95% of cases prior to March 7 (**Figure-1B, Appendix-1 Table-1**). By May 25, 41.6% of cumulative cases were diagnosed in congregate settings and 58.4% were diagnosed outside congregate settings, including 3.8% travel-related, 37.8% in community settings (18.5% with or 19.3% without an epidemiological link or close contact), and16.8% with missing information (**Figure-1B, Appendix-1 Table-1**). Proportions of daily new diagnoses by setting over time are shown in **Appendix-1 Figure-2B and Appendix-1 Table-1:** on May 25^th^, 15.4% of new diagnoses were in congregate settings, 0.6% were travel-related, 76.8% were in community settings, and 7.3% with missing information.

**Figure 1.**
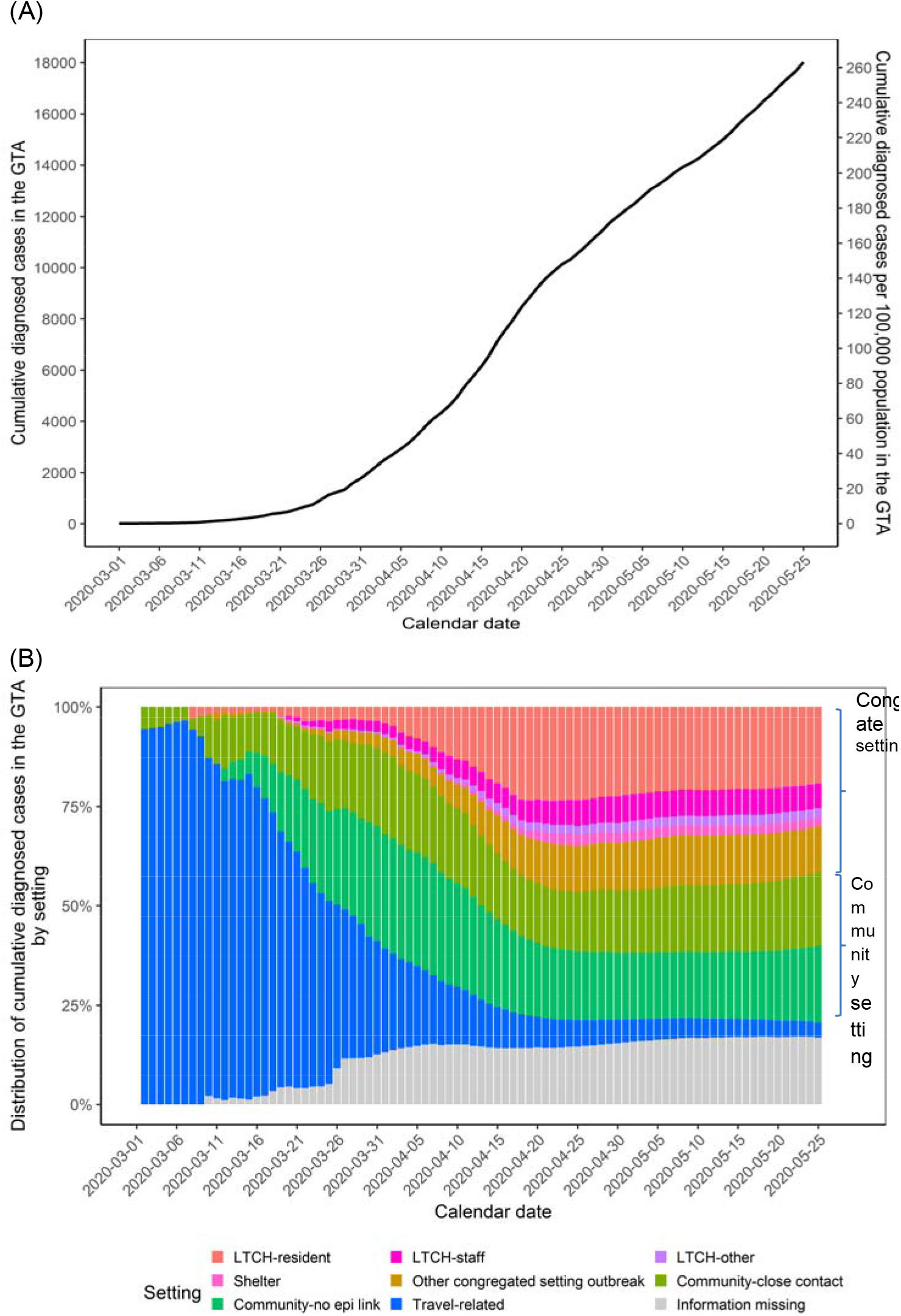
The (A) total number and (B) distribution of cumulative diagnosed COVID-19 cases in the Greater Toronto Area by outbreak setting over time. Settings are defined as mutually exclusive categories by the order shown in the graph (from top to bottom) in the event of multiple exposures. LTCH-other may include volunteers; other congregate outbreak settings include hospitals, correctional facilities, retirement homes, group homes, and other not yet classified such as workplaces. Information missing category excludes congregate setting. The calendar date refers to the date the case was reported to the public health unit. Data sources: iPHIS, the integrated Public Health Information System. Abbreviations: GTA, Greater Toronto Area; LTCH, long-term care homes.

In March, diagnoses transitioned from predominantly travel-related to community settings. By the end of March, 28.5% of cumulative cases were travel-related, 48.6% were in community settings, and 10.3% were in the congregate outbreak settings (**Figure-1B**). A sharp increase in cases in congregate settings, particularly among LTCH residents followed in April. From April 1 to April 20, the proportion of cumulative cases increased in each congregate setting: LTCH residents (from 4.1% to 23.3%); LTCH staff (from 2.8% to 5.7%); persons using shelters (from 0% to 2.4%); and in other congregate settings (from 4.0% to 10.9%). The cumulative proportion of cases in congregate settings remained relatively stable thereafter (**Figure-1B**).

Of all cases in congregate settings by May 25, nearly half (46.5%) were among LTCH residents, and 5.4% were among persons using shelters (**Appendix-1 Figure-3**). **Appendix-1 Figure-3 and Appendix-1 Table-1** show the distribution and number of diagnoses by type of congregate setting. The sex and age distribution of cases among LTCH residents, in shelters, and among the rest of the population varied considerably (**Table-1**). Compared to the diagnosed cases in the rest of the population, LTCH residents diagnosed with COVID-19 were older (74.7% aged 80 years or more vs. 7.2%, p<0.001) and more likely to be a female (66.4% vs. 53.2%, p<0.001); while persons using shelters and diagnosed with COVID-19 were younger (71.3% aged less than 50 years vs. 52.4%, p<0.001) and less likely to be a female (41.6% vs. 53.2%, p<0.001) (**Table-1**).

**Table 1.** **Comparison across outbreak settings in the Greater Toronto Area regarding the cumulative risk of diagnosis, testing and case fatality of COVID-19 by May 25th, 2020**.

| Measures | LTCH residents | Persons using shelters | The rest of the population | P-value** |
| --- | --- | --- | --- | --- |
| Population size* | 28316 | 10588 | 6808890 |  |
| Number of diagnosed cases, overall | 3485 | 402 | 14133 |  |
| Female, N(%)*** | 2239 (66.4%) | 165 (41.6%) | 7473 (53.2%) | <0.001 |
| Age, years, N(%)*** |  |  |  | <0.001 |
| <50 | 20 (0.6%) | 286 (71.3%) | 7411 (52.4%) |  |
| 50-59 | 50 (1.4%) | 70 (17.5%) | 2972 (21.0%) |  |
| 60-69 | 197 (5.7%) | 27 (6.7%) | 1917 (13.6%) |  |
| 70-79 | 613 (17.6%) | 14 (3.5%) | 818 (5.8%) |  |
| 80+ | 2605 (74.7%) | 4 (1.0%) | 1015 (7.2%) |  |
| Number of individuals tested for COVID-19, overall | 21857 | 1477 | 163476 |  |
| Female, N(%) | 14970 (68.5%) | 420 (28.4%) | 100629 (61.6%) | <0.001 |
| Age, years, N(%) |  |  |  | <0.001 |
| <50 | 171 (0.8%) | 838 (56.7%) | 83846 (51.3%) |  |
| 50-59 | 519 (2.4%) | 320 (21.7%) | 30797 (18.8%) |  |
| 60-69 | 1613 (7.4%) | 208 (14.1%) | 20399 (12.5%) |  |
| 70-79 | 3624 (16.6%) | 76 (5.1%) | 11502 (7%) |  |
| 80+ | 15930 (72.9%) | 35 (2.4%) | 16932 (10.4%) |  |
| Number of deaths among diagnosed cases, overall | 915 | 3 | 506 |  |
| Female, N(%) | 530 (59.7%) | 0 (0%) | 210 (41.5%) | <0.001 |
| Age, years, N(%) |  |  |  | <0.001 |
| <50 | 0 (0%) | 0 (0%) | 21 (4.2%) |  |
| 50-59 | 7 (0.8%) | 2 (66.7%) | 40 (7.9%) |  |
| 60-69 | 35 (3.8%) | 0 (0%) | 80 (15.8%) |  |
| 70-79 | 132 (14.4%) | 1 (33.3%) | 118 (23.3%) |  |
| 80+ | 741 (81%) | 0 (0%) | 247 (48.8%) |  |
| Diagnosed cases per 100,000 |  |  |  |  |
| Absolute value | 12308 | 3797 | 208 |  |
| Relative value | 59.2 | 18.3 | Reference |  |
| Proportion of population tested for COVID-19, % |  |  |  |  |
| Absolute value | 77.2 | 13.9 | 2.4 |  |
| Relative value | 32.2 | 5.8 | Reference |  |
| Proportion of tests with positive results, % |  |  |  |  |
| Absolute value | 15.3 | 4.8 | 6.7 |  |
| Relative value | 2.3 | 0.7 | Reference |  |
| Age- and sex-adjusted test positivity rate ratio (95% CI)**** | 2.5 (2.3-2.8) | 0.8 (0.5-1.1) | Reference |  |
| P-value | < 0.001 | 0.036 | Reference |  |
| Case fatality proportion |  |  |  |  |
| Absolute value | 26.3 | 0.7 | 3.6 |  |
| Relative value | 7.3 | 0.2 | Reference |  |
| Age- and sex-adjusted case fatality rate ratio (95% CI)**** | 1.4 (1.1-1.9) | 0.6 (0 - 3.3) | Reference |  |
| P-value | 0.028 | 0.65 | Reference |  |

Abbreviations: LTCH, long-term care home; CI: confidence intervals.
\*LTCH residents population size approximated by the total LTCH bed capacity in the Greater Toronto Area; persons using shelters population size approximated by the estimated number of people experiencing homelessness in the Greater Toronto Area (Appendix-2); the rest of the population size estimated by the total census population size of subtracting population size of LTCH residents and persons using shelters.
\*\*Comparison using Chi-squared tests.
\*\*\*Of 18020 diagnosed cases, 206 had unknown sex, and 1 had unknown age; age and sex distribution proportions based on non-missing information.
\*\*\*\*Estimated using quasipoisson regression models, adjusting for age and sex.

### Cumulative diagnoses per capita by setting

Cumulative diagnoses per capita by setting over time are shown in **Figure-2**. Cumulative diagnosed cases per 100,000 population were 59-fold and 18-fold higher among LTCH residents (12308), and persons using shelters (3797), respectively, compared to the rest of the population (208) (**Table-1**).

**Figure 2.**
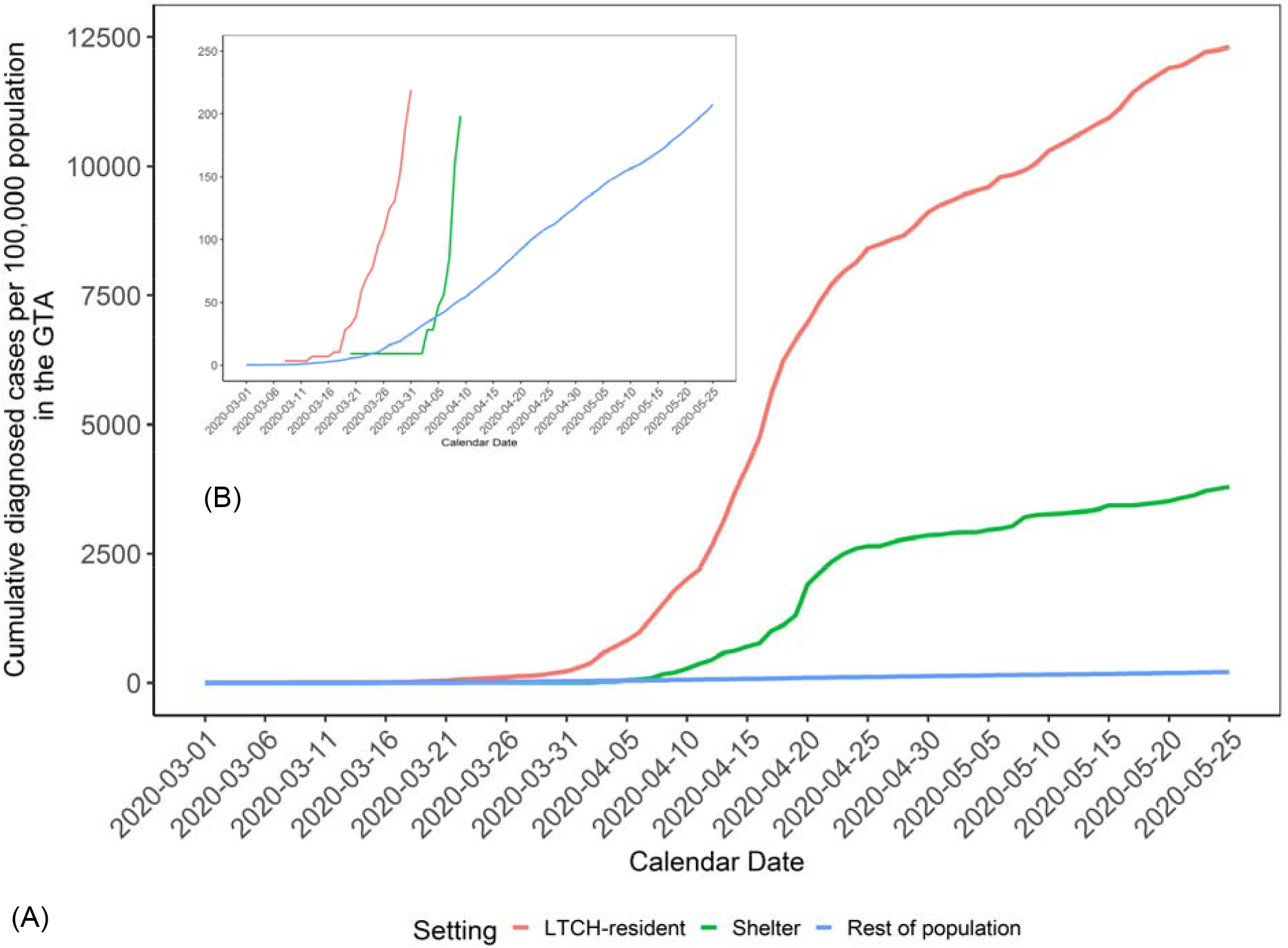
(A) Comparison of cumulative diagnosed cases per capita over time by outbreak setting in the Greater Toronto Area. (B) has the same information as (A) but with different y-axis range. The calendar date refers to the date the case was reported to the public health unit. Data sources: iPHIS, the integrated Public Health Information System. Abbreviations: GTA, Greater Toronto Area; LTCH, long-term care homes.

### Per-capita testing volume and positivity rate

By May 25, 77.2% of LTCH residents and 13.9% of persons using shelters had been tested at least once, compared to 2.4% of the rest of the population (**Table-1**;**Appendix-1 Figure-4**). Cumulative test positivity was: 15.3% (LTCH residents), 4.8% (persons using shelters), and 6.7% (rest of the population). Among those tested, the age and sex-adjusted test positivity rate ratio was 2.5 (95% CI: 2.3-2.8) among LTCH residents, and 0.8 (95% CI: 0.5-1.1) among persons using shelters, compared to the rest of the population (**Table-1; Figure-3A**).

LTCH residents’ test positivity changed over time with varying testing volume: the daily new testing positivity proportion spiked in early April, with 20%-60% of LTCH residents testing positive each day (**Figure-3B**). After April 20, and as per-capita testing volumes rose, test positivity among LTCH residents fell to 10%, similar to positivity in the rest of the population (**Figure-3B**). Test positivity among LTCH residents re-spiked in mid-May with fewer tests (**Figure-3B**).

**Figure 3.**
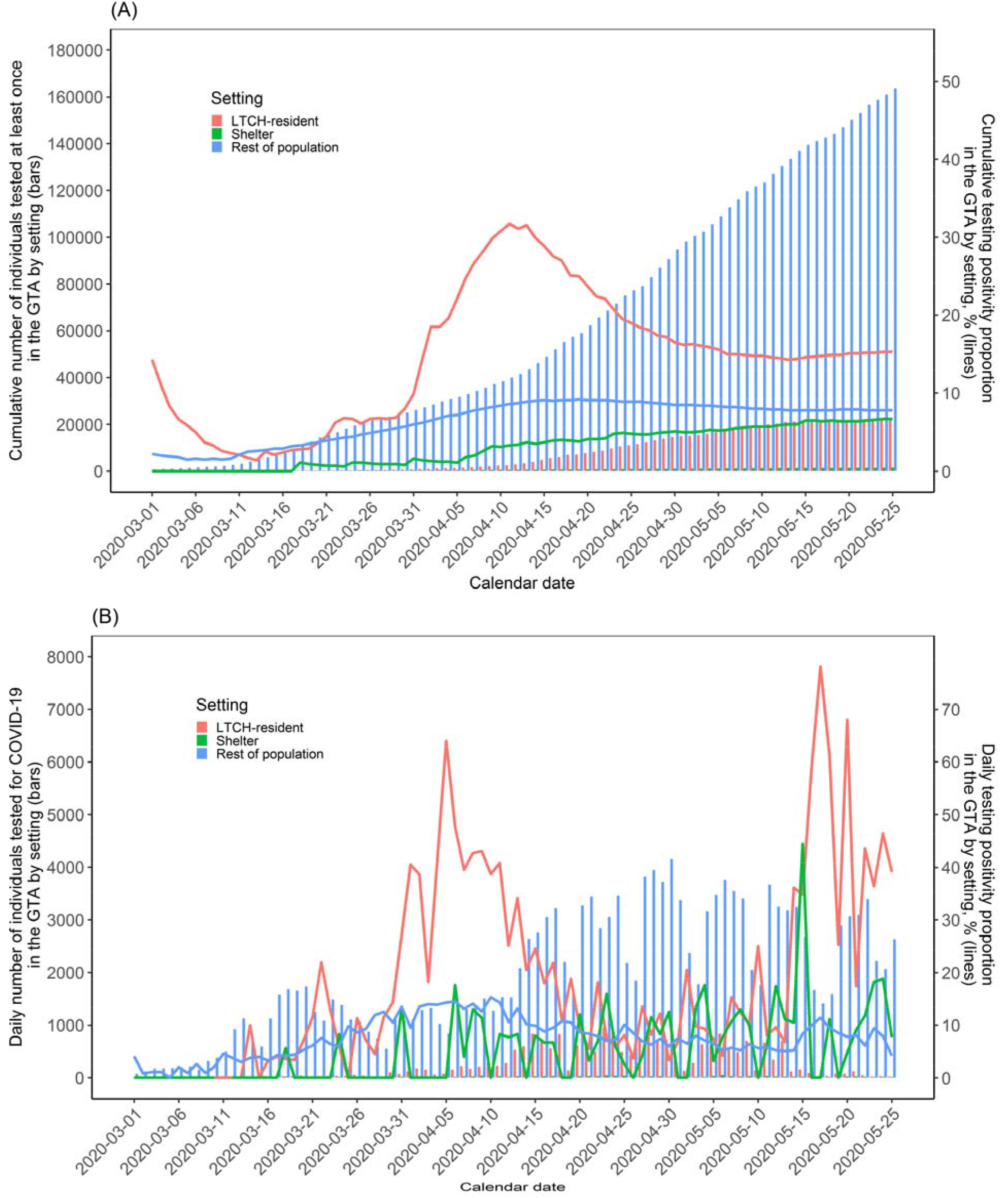
Cumulative (A) and daily (B) number of individuals tested for COVID-19 and test positivity proportion over time by outbreak setting in the Greater Toronto Area. The calendar date refers to the date when specimen was collected. Data sources: OLIS, the Ontario Laboratories Information System. Abbreviations: GTA, Greater Toronto Area; LTCH, long-term care homes.

### Case fatality proportion

As of May 25, 915 (64.3%), 3 (0.2%) and 506 (35.5%) deaths were reported among LTCH residents, persons using shelters, and the rest of the population, reflecting a case fatality proportion of 26.3%, 0.7%, and 3.6%, respectively (**Table-1**). The age- and sex-adjusted case fatality rate was 1.4 times (95% CI: 1.1-1.9) higher among LTCH residents compared with the rest of the population (**Table-1**).

## Interpretation

Diagnosed cases of COVID-19 in the GTA have shifted from travel-related, through the community, and are now divided between outbreaks in congregate settings such as LTCH and shelters and community settings. Congregate settings represent a disproportionate risk of cases, and in the context of LTCH residents, higher case-fatality among those diagnosed.

The time-course of the micro-epidemics within a large city such as the GTA raise questions about how transmission may have moved through physical (and thus, social) networks defined along intersections of architectural, occupation, and socio-economic factors. LTCH were quick (on March 14^th^, 2020) to close visitations (37-39) and thus connections between residents and the wider community were largely limited to LTCH staff and volunteers. Meanwhile, early (in early March) efforts to implement enhanced infection control practices, screening, triage, and a temporary housing strategy for persons experiencing homelessness and who were awaiting test results may also have delayed the onset of outbreaks in shelters (40, 41). Neighborhood-level measures of marginalization suggest that it also took time for the virus to spread via travel in the context of higher-income social and physical networks than through lower socio-economic networks and households where physical distancing may be limited by high density-dwellings (42-44). Some community cases may reflect close contacts or an epidemiological link with congregate settings; for example, as members of households of persons who work or volunteer in facilities and essential workplaces (45, 46). Thus, alongside the overall reductions in outdoor and between-household contacts in the community (47), the overall GTA epidemic may have now concentrated in congregate settings and in community households, with additional work needed to discern connections between networks.

The size and trajectory in the per-capita rate of diagnosed among LTCH residents and among persons using shelters could reflect underlying differences in testing and/or to differential vulnerabilities to outbreaks. After the initial LTCH outbreaks were detected in late March, Ontario instituted a wider scope for testing in LTCH – including asymptomatic residents (48) – which led to a surge in cases identified through this point-prevalence approach to outbreak investigations (49). Declines thereafter in the LTHC residents’ positivity rate may reflect a combination of: infections averted; surge in testing already diagnosed the most vulnerable; an effective shield or herd immunity achieved within facilities (deaths among existing residents and without new admissions means fewer susceptible residents for an outbreak to persist) (50). The higher positivity rate in late May among LTCH residents with a smaller tested proportion may suggest more targeted testing during this latter period.

The 2.5-fold higher cumulative positivity among LTCH residents, after adjustment for age and sex, suggests a combination of: higher risk transmission environments or differences in testing practices and criteria for LTCH residents versus the rest of the GTA population. For example, observed cases may reflect a higher proportion of asymptomatic infection among LTCH residents, because until May 24^th^, the criteria for testing outside the context of congregate settings were more risk-based (symptoms, epidemiological link or close contact/exposures) (51-53).

Finally, death among LTCH residents diagnosed with COVID-19 accounted for approximately two thirds of deaths among all diagnosed cases in the GTA, similar to Ontario overall, British Columbia, and Québec (54-56). A higher age- and sex-adjusted case fatality rate among LTCH residents as compared with the rest of the population may reflect underlying differences in comorbidities associated with COVID-attributable mortality and/or goals of care (57). Our findings suggest that the 13-fold higher COVID-related mortality among LTCH residents as compared with all non-LTCH elders in Ontario (58), could stem from both the higher risk of infection and rates of COVID-19 diagnoses, and a higher case fatality among LTCH residents diagnosed with COVID-19.

There are important limitations to note. First, we were limited to quantifying disproportionate risks to sub-populations on whom we could obtain data on the population size denominators (as such, e.g., we could not estimate diagnoses per capita for LTCH staff and for retirement homes). Future work in epidemic appraisal necessitates population size and per-capita estimates across each type of outbreak setting, and additional disaggregation that are now being collected (ethnicity, social-economic status, comorbidities) as part of the person-level data (59-61). Second, ‘rest of the population’ subsumed other congregate facilities, and thus our estimates of the relative difference in per-capita testing and positivity may be an underestimate as other congregate facilities are known to be associated with more testing (e.g. hospitals (52, 53) and risk of outbreaks (62)). Third, we assumed that the population denominators for each outbreak setting were mutually exclusive and static. However, during the course of epidemic, there could be shifts in setting-specific population size (e.g., LTCH residents may have moved to home;. although new admissions to LTCH were halted during the outbreak (63), there was an active strategy to support housing at LTCH for older adults who were underhoused (64). Fourth, the large underestimate in the OLIS data, of persons experiencing homelessness and diagnosed with COVID-19 (**Appendix-1 Figure-1C**) suggests the proportion tested and test positivity among persons experiencing homelessness may be a large underestimate, and thus must be interpreted with caution.

Heterogeneity across micro-epidemics signal the need for setting-specific and population-specific strategies in the next phase of the public health response in Canada, which could be guided by next steps of work including modeling the risks of onward transmission across each layer of heterogeneity and connections between networks.

## Data Availability

The findings of this study are supported by 1) data from the Public Health Ontario (PHO) integrated Public Health Information System (iPHIS), 2) the Ontario Ministry of Long-Term Care Long-Term Care Homes Tracker; and 3) the Ontario Laboratories Information System (OLIS). The iPHIS and Long-Term Care Homes Tracker were accessed via the Ontario COVID-19 Modelling Consensus Table. The OLIS data were accessed via ICES. In addition, data that are sourced publicly are made available in the paper as appendix or as per the referenced links.

## Acknowledgements

Reported COVID-19 cases were obtained from the Public Health Ontario (PHO) integrated Public Health Information System (iPHIS), via the Ontario COVID-19 Modelling Consensus Table, and with approval from the University of Toronto Health Sciences REB (protocol No. 39253). This Ontario Laboratories Information System (OLIS) data and linkages were supported by ICES, which is funded by an annual grant from the Ontario Ministry of Health and Long-Term Care (MOHLTC). Parts of this material are based on data and information compiled and provided by MOHLTC and the Canadian Institute for Health Information (CIHI). The analyses, conclusions, opinions and statements expressed herein are solely those of the authors and do not reflect those of the funding or data sources; no endorsement is intended or should be inferred.

